# Convolution Neural Networks for Point-of-Care Diagnostics of Bacterial Infections in Blood

**DOI:** 10.1101/2022.01.03.22268712

**Authors:** Omkar Hegde, Ritika Chatterjee, Durbar Roy, Vivek Jaiswal, Dipshikha Chakravortty, Saptarshi Basu

**Author notes:** Corresponding Author email –. These authors contributed equally as first authors (Omkar Hegde, Ritika Chatterjee). These authors contributed equally as second authors (Durbar Roy, Vivek Jaiswal).

## Abstract

A droplet of blood, when evaporated on a surface, leaves dried residue—the fractal patterns formed on the dried residues can act as markers for infection present in the blood. Exploiting the unique patterns found in the residues of a naturally dried droplet of blood, we propose a Point-of-Care (POC) diagnostic tool for detecting broad-spectrum of bacterial infections (such as *Enterobacter aerogenes, Staphylococcus aureus, Klebsiella pneumoniae, Acinetobacter baumannii, and Pseudomonas aeruginosa, Salmonella Typhi*) in blood. The diagnosis process we propose is straightforward and can be performed with the following steps: A droplet of blood (healthy or infected) of volume range 0.5 to 2 *μl* is allowed to dry on a clean glass surface and is imaged using a conventional optical microscope. A computer algorithm based on the framework of convolution neural network (CNN) is used to classify the captured images of dried blood droplets according to the bacterial infection. In total, our multiclass model reports an accuracy of 92% for detecting six bacterial species infections in the blood (with control being the uninfected or healthy blood). The high accuracy of detecting bacteria in the blood reported in this article is commensurate with the standard bacteriological tests. Thus, this article presents a proof-of-concept of a potential futuristic tool for a rapid and low-cost diagnosis of bacterial infection in the blood.

## INTRODUCTION

Antimicrobial resistance (AMR) has become a cause of global concern, aggravated by a lack of data on the different mechanisms and factors causing its emergence and spread. According to a WHO report, bacterial infections alone cause nearly 1 million deaths per year worldwide; this number is estimated to increase to 10 million per year by 2050, owing to AMR^1^. Entero-pathogens (such as *Salmonella* Typhi) and ESKAPE (*Enterococcus faecium, Staphylococcus aureus, Klebsiella pneumoniae, Acinetobacter baumannii, Pseudomonas aeruginosa, and Enterobacter aerogenes*) pathogens are the deadliest, and every passing year they cause millions of infections and thousands of deaths worldwide^2–6^. These bacteria are clinically relevant strains causing ∼42% bloodstream infection and have been associated with significant mortality and morbidity^7,8^. The WHO report lists these pathogens as priority antibiotic-resistant pathogens as they are associated with a significant amount of mortality and morbidity due to the emergence of multiple drug resistance (MDR) and extensive drug resistance (XDR) strain^6^. Even developed economies like the United States^9^ and other European countries^10^ are finding it difficult to cope with the increasing AMR; an estimated cost of billions of dollars is attributed to the health care expenditure of hospital-acquired and community-acquired AMR infections^9^. The first step to stop the spreading of infection would be to diagnose it. Timely diagnosis of the causative organism and subsequent treatment of bacterial infection can save millions of lives and inhibit the generation of various drug-resistant strains. “Test, isolate (the person infected), and treat” is the primary formula used by advanced countries to stop the spread of infection, as evidenced during the Global COVID-19 pandemic^11^. Thus, it is crucial to test rapidly to prevent the further spread of infection and any possible outbreak.

Since the abovementioned bacterial strains (Enteropathogens and ESKAPE pathogens) majorly cause bloodstream infections^12,13^ and can mainspring changes in the composition and fluid properties of the blood, it is prudent to test blood for diagnosis of these infections. Moreover, blood profile indicates the overall health of the individuals; thus, it is a general practice to test blood for several abnormalities, including bacterial infections. Conventionally, various tests and investigations are undertaken to diagnose diseases in blood—such as a complete blood count, chemistry profiling of urea and electrolytes, level of C-Reactive Protein (CRP), glucose, and coagulation screen, to name a few. Several biomarkers have been reported in the literature to diagnose sepsis^14^, such as pro-inflammatory and anti-inflammatory cytokines and chemokines, altered cell surface marker of immune cells. One of the main drawbacks of these markers is that they have low specificity and high cost associated with high-end technique requirements for the detection^15^. The gold standard for bacterial infection diagnosis is bacteriological culture and staining^16^. Infectious agents are isolated and identified using various selective and differential culture media. However, bacteriological culture diagnosis of the infection is often time-consuming (require up to 48 hours). Alternatively, polymerase chain reaction (PCR) based diagnostics are faster than standard bacteriological culture-based methods. Nonetheless, due to high sensitivity, it often can give false-positive results. Another major disadvantage for PCR-based detection involves too high cost and specialized instrumentation and setup requirement, which may not be feasible in semi-urban or rural areas^17–20^. Therefore, there is an urgent need for innovative solutions to create accurate, low-cost, rapid diagnostic tools to detect bacterial infections in the blood. This article addresses the above problems by providing a simple method to rapidly detect a broad spectrum of bacterial infections in the blood (with a turnaround time of less than 15 minutes) by leveraging the unique patterns formed on the residue of the dried blood droplets.

With the advent of fundamental research on droplet evaporation and colloidal self-assembly in droplets in the past few decades, the dried precipitates of biological fluids have been deeply studied^21–30^. A few of these studies have found applications in biomedical diagnostics^31–34^. The competitive interaction of multi-component elements in biological fluid like blood leads to the formation of rich patterns on the dried droplet precipitate^35^ such as distinctive radial, spiral, and tangential cracks, color variations, varying thickness of the coffee rings, finger formations, etc. (Refer to image S1 in the supplementary information for the image of a dried droplet of blood taken from a healthy volunteer). These features depend on the properties of the fluid (and thereby the components present in it), which in turn can be related to the health of a person. Notably, in blood droplets, it has been shown previously that the flow inside the droplet is driven by Marangoni, wettability, and natural evaporation leads to the formation of distinctive patterns on the dried residues of the drop for anemic and hyperlipidemic blood samples^36–38^. Besides, the final dried blood droplet pattern is susceptible to drying conditions such as temperature^39^ and humidity^40^, placement of blood droplet^41^ (gentle or with impact), and depends on several other factors such as blood groups^42^, ethnicity^43^, age^44^, gender^43^, size, shape, and mobility of cells^45^. Further, blood infection leads to cellular changes such as Leucocytosis^63^ or leukopenia, and biochemical changes include markers released upon red blood cell lysis or white blood cell lysis^63^. Thus, the blood carrying the infection would also exhibit different properties (compared to the healthy blood), leading to a unique pattern formation on the dried blood drop^46^; for example, it is possible to detect malaria in dried blood droplets using a simple light microscope imageing^47^.

However, the differences in the final pattern on the blood droplet residue could be due to several reasons, and markers for an infection in these patterns can be subtle. Thus, the human interpretation of optical images of dried blood droplets for disease diagnosis can be prone to errors; it is inevitable to use computer-assisted diagnosis. Also, the use of computers for the diagnosis will lead to a rapid and more accurate diagnosis. Bio-Medical diagnostics have extensively used Deep learning (DL) and Convolutional Neural networks (CNN). For example, with the help of images generated by CT scans, breast cancer, lung nodule, and segmentation can be detected using CNN and DL^19-27^. These computer models reported 63-85% accuracy (AlexNet 63.98% ^28^, DenseNet 80.7% ^29,^ and Wang et al. 85% ^30^), but the training data set had very few runs (less than five runs). Recently, Machine Learning (ML) analysis was used to discriminate dried blood droplet patterns with varying physiological conditions, i.e., changes in dried blood droplet patterns before or after physical exercise of the volunteers were detected using ML with a 95% prediction accuracy^48^. This demonstrates that ML is a potent tool to classify patterns formed by blood droplets with even slight changes in blood chemistry induced by the physical exercise of volunteers. However, it remains unclear if the ML or DL can detect a wide variety of bacterial pathogens in the blood.

The current article presents a proof-of-concept of an alternative low-cost and rapid point-of-care diagnostic tool by imaging a dried droplet of blood using simple optical microscopy. It has been demonstrated that a wide range of deadly pathogens such as (*Enterobacter aerogenes, Staphylococcus aureus, Klebsiella pneumoniae, Acinetobacter baumannii, and Pseudomonas aeruginosa, Salmonella Typhi*) present in the blood can be detected using the framework of CNN. The diagnostic procedure is described as follows: An image of a dried blood droplet (that may contain the target infection) on a glass substrate is captured using an optical microscope. The captured image is fed into an in-house developed computer algorithm that predicts the possible infection based on CNN architecture. Our model reports an accuracy of 92% for distinguishing the infection of the abovementioned bacterial species in the blood. The computer algorithm for this application is developed by training and testing the algorithm in a supervised learning environment with a large number of images of blood droplets (∼35,000 images), whose status of infection is known apriori. The model accuracy remains invariant over the large dataset.

## RESULTS AND DISCUSSION

Blood collection from the volunteers is detailed in the experimental procedure section. The blood is spiked with bacteria in-*vitro* (individual bacterial species are spiked in different vials of blood (Fig.1 (a) and (b)) - *Enterobacter aerogenes* (EA), *Staphylococcus aureus* (SA), *Klebsiella pneumoniae* (KP), *Acinetobacter baumannii* (AB), *Pseudomonas aeruginosa* (PA)), *Salmonella* Typhi (ST)). The unspiked blood sample is used as a control (C) against the abovementioned spiked samples, i.e., it does not contain any bacteria and is considered a healthy blood sample. These samples are incubated for 24h at 37°C. Each blood sample in the vials (including the control) is considered an individual class and labeled accordingly (see Fig. 1(b)). Several hundred droplets (of volume 0.5 to 2 μl) of blood samples (spiked/unspiked) are deposited on a clean glass slide on a given day and allowed to evaporate in the controlled atmospheric condition (26 ±5 °C and 45 ±10% RH) (Fig.1 (c) and (d)). The droplets are imaged using a conventional optical microscope—a total of 35000 images are acquired (number of images in each class (∼5000 images of each class). Sample images of the dried blood droplet of the glass slide spiked with different bacteria, and the control (unspiked) is shown in Fig.1 (e). The blood droplet evaporates in constant contact radius (CCR) mode as the droplet remains pinned throughout evaporation (irrespective of any class). The droplets generally assume the shape of a spherical cap as the Bond number (Bo) is <<1 and the dried residue is of a circular shape. However, few droplets show a shape disparity due to errors in dispensing the drop from the micropipette. Thus, amongst the nearly 47,000 images taken, about 13,000 images are discarded to disregard the shape disparity of the droplet. The analysis in this article is based on 34469 images considered after filtration. It is evident from Fig.1 (e) that it is difficult to classify the blood droplet residues according to the bacteria present in them from a visual inspection of the images. Hence, we have developed an image classification computer algorithm using CNN to recognize different classes.

**Figure 1.**
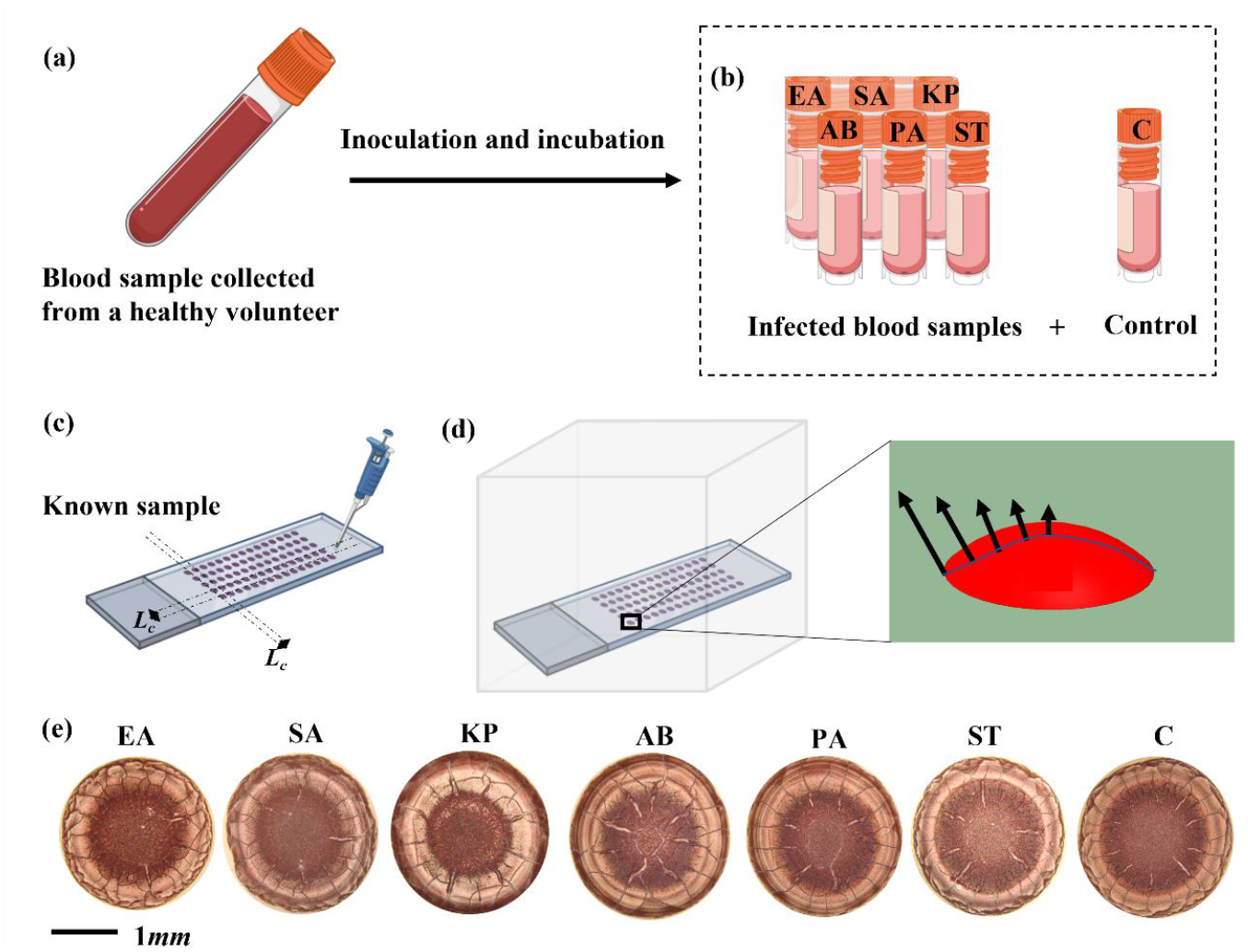
**Evaporation triggered pattern formation in infected and healthy blood droplets. (a) 15 ml of blood is collected from a healthy volunteer in vacutainer tubes. (b) the collected blood is infected with bacteria (in-*vitro*) and is labeled as follows: *Enterobacter aerogenes* - EA, *Staphylococcus aureus* - SA, *Klebsiella pneumoniae* - KP, *Acinetobacter baumannii* - AB, *Pseudomonas aeruginosa* - PA, *Salmonella* Typhi -ST. The blood sample that does not contain any infection is considered as a control and is labeled as - C. All the samples are incubated for 24h at 37° C. The individual labels are referred to as “class”. (c) A few hundreds of droplets of 0.5-2 μl of a known sample (as labeled in (b)) are gently placed on clean glass slides using a micro-pipette. The individual droplets are placed far apart (*Lc*) such that there is no vapor-mediated interaction between them. (d) The deposited droplets are allowed to evaporate in an enclosed chamber. The evaporation profile is shown in the zoomed-in image of the droplet, with the evaporation flux being maximum at the edge and minimum at the center of the drop. (e) Sample images of dried blood droplets labeled according to their class (the respective label is on top of the image). The label at the top of the image indicates the species of bacterial infection in the blood. The last image in the row is not infected with bacteria and thus is the control.**

The algorithm is developed in Python programming language and uses seven libraries to analyze the data set: NumPy^49^, Keras^50^, Matplotlib^51^, Seaborn^52^, Scikit^53^, Tensorflow^54^, and OpenCV^55^ (Refer to Fig. 2(a), Matplotlib, Seaborn are for plotting the data). The data set (captured images of blood droplets) is imported and labeled according to the given class. Prior to analysis, the image data set is segregated in the ratio of 80:20 for training and testing, respectively (see Fig. 2(b)). This segregation is done randomly to avoid any bias. The images are further pre-processed using a series of operations using the ImageDataGenerator function in the Keras library to maintain uniformity for variables like contrast, color, and positioning of the drop in the image. The processed images are discretized and equalized into an array of numbers (vector). However, analyzing this data on traditional computing infrastructure (the hardware used is Intel(R) Core i7 2.90 GHz 64 GB RAM) is a challenge as the volume of data generated by the images is very large.

**Figure 2.**
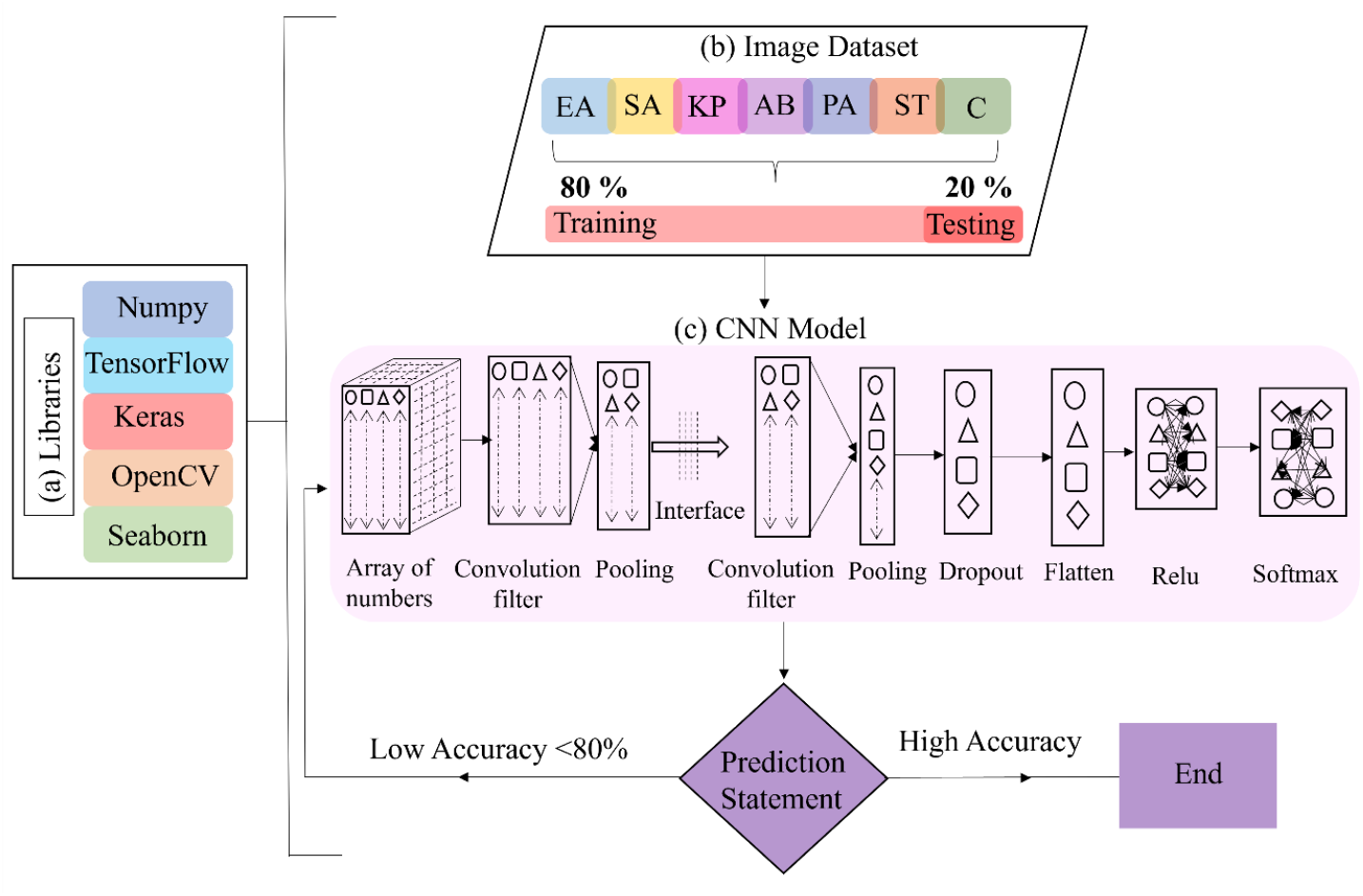
**The overall design of the CNN algorithm used to train the computer to classify images of the residues of dried blood droplets according to their classes. (a) The algorithm imports standard libraries used for CNN. Importantly, NumPy^49^, Keras^50^, Tensorflow^54^, OpenCV^55^, and Seaborn^52^ are imported libraries, to name a few. (b) The imported image data set consisting of ∼35000 images are labeled as per their respective classes. The image dataset is divided into the training and testing sets in the ratio 80:20, respectively. The model is trained using the training data set and is validated using the test dataset. The test dataset is not used during training, thus ensuring no data leakage from the test dataset. (c) The image data set is converted to a number array (vector) and analyzed using multiple layers. Model parameters such as epochs, kernel size for convolutional layers, image size, and learning rate are fed into the model, and the model is trained using the training set. The model makes a prediction using the testing dataset. Iterations are repeated with a new set of values for model parameters until the highest possible accuracy is achieved.**

Hence the algorithm is developed in a two-step process. First, the model is optimized for binary classes, i.e., the algorithm is trained to distinguish between any two classes with high accuracy. Since there are seven classes, the model for ^7^C_2_ binary classes is optimized (see Fig. 3). Second, the parameters used for optimizing the binary classes (such as epochs - the number of times the algorithm iterates the whole dataset for training, kernel size for convolutional layers^51^, image size, and learning rate - the variation in the model with updated weights) are used in the multiclass model with all the classes included, as shown in Fig. 3 (7 classes including the control case). While the basic structure of the algorithm remains the same, the parameters optimized for binary classes are used as a template for the multiclass model with all the classes.

**Figure 3.**
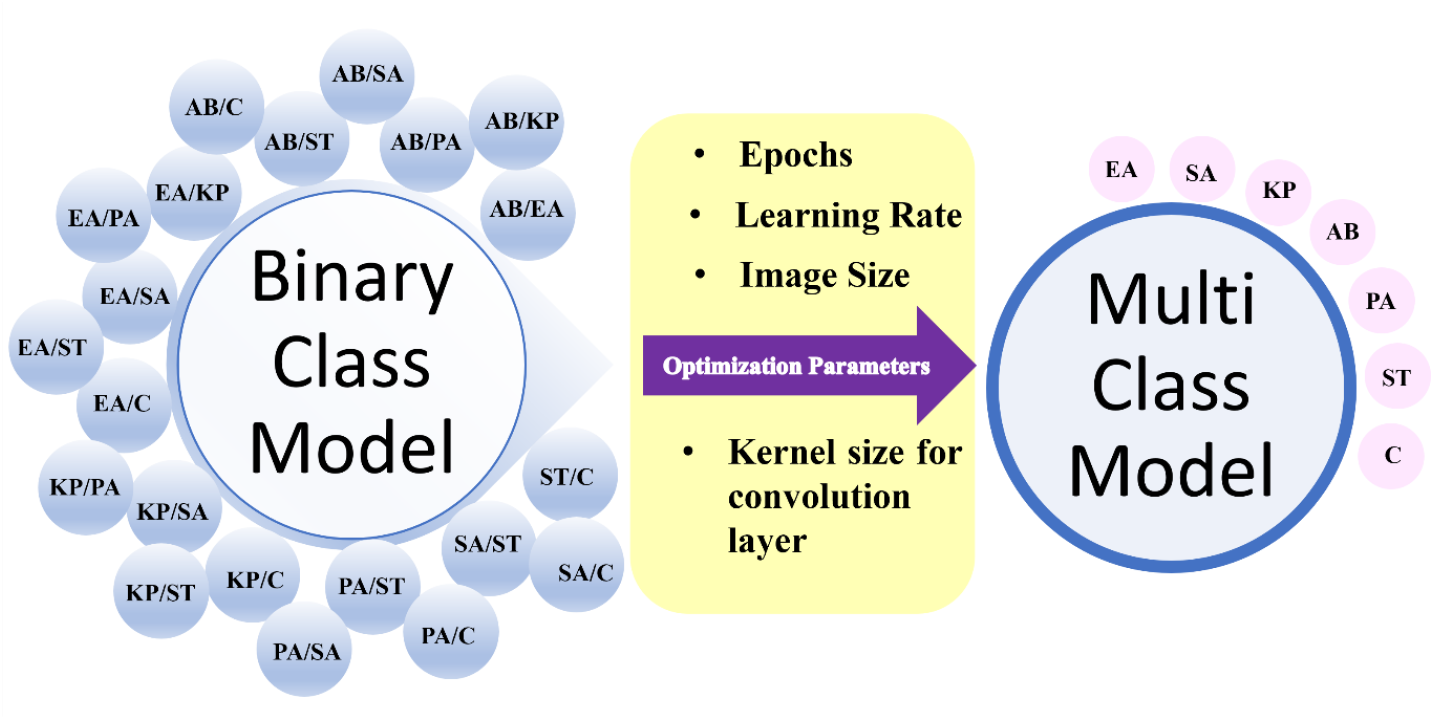
**Schematic representation of the two-step development of the model used in the article. Optimal model parameters such as the epoch, learning rate, image size, and kernel size for the convolution layer obtained from the ^7^C_2_ binary classes are adopted into the multiclass model consisting of seven classes.**

To examine the algorithm for its ability to distinguish between the binary classes, the vector output of the processed images from any two classes is input into the CNN Model. The CNN model consists of the following layers: convolution layer^51^, pooling layer^56^, dropout layer^52^, flattening layer^53,^ and fully connected layer^54^ (Refer to Figure 2(c)). The modeling parameters such as filters used and kernel size in the convolution layer, the prevention of overfitting the data in the dropout layer, and units in the dense layer^50^ have a crucial role in improving the overall accuracy of detection. The values of epochs, learning rate, kernel and image size for which the highest accuracy of prediction is achieved are considered optimized values. Using the trial and error method, the optimization of the model (for the given vector) is achieved by setting the values of epochs, learning rate, kernel size, and image size - 50, 10^−4^, 9×9, and 256 pixels, respectively. The trained model is tested using the test image data set (20 % of the segregated image data set for testing) and the results of accuracy, precision, and recall for 21 binary classes are given in table 1.

**Table 1:** Accuracy, precision, and recall values for ^7^C_2_ Binary classes.

| Binary class | Accuracy | Precision | Recall |
| --- | --- | --- | --- |
| AB/ EA | 0.97 | 0.97 | 0.96 |
| AB/KP | 0.84 | 0.96 | 0.92 |
| AB/PA | 0.89 | 0.96 | 0.95 |
| AB/SA | 0.91 | 0.95 | 0.93 |
| AB/ST | 0.92 | 0.99 | 0.98 |
| AB/C | 0.85 | 0.91 | 0.92 |
| EA/ KP | 0.98 | 0.98 | 0.98 |
| EA/PA | 0.88 | 0.95 | 0.96 |
| EA/SA | 0.92 | 0.92 | 0.92 |
| EA/ST | 0.94 | 0.98 | 0.97 |
| EA/C | 0.98 | 0.98 | 0.98 |
| KP/PA | 0.92 | 0.93 | 0.92 |
| KP/SA | 0.91 | 0.9 | 0.92 |
| KP/ST | 0.93 | 0.94 | 0.94 |
| KP/C | 0.80 | 0.85 | 0.81 |
| PA/SA | 0.96 | 0.96 | 0.96 |
| PA/ST | 0.92 | 0.98 | 0.97 |
| PA/C | 0.92 | 0.94 | 0.93 |
| SA/ST | 0.98 | 0.98 | 0.98 |
| SA/C | 0.88 | 0.9 | 0.88 |
| ST/C | 0.92 | 0.93 | 0.92 |

The performance of the model is gauged importantly by the following output parameters: accuracy, loss, precision, and recall. The accuracy of the model is the ratio of correctly predicted to the total samples. The loss is the summation of errors made in prediction by the model on the training and test image datasets (see Fig. 5 (b)). The precision is the percentage of actual positive samples to the predicted positive samples (which may be correct or incorrect). Recall (sensitivity) is the ratio of correctly predicted positive samples out of total positive samples. As shown in Table 1 (for binary classes), the accuracy, precision, and recall range from 80-98% to 85-98% and 81-98%, respectively. The model parameters used for the binary classes are considered to be optimized as the accuracy achieved for binary classes is maximum for the given image data set. The optimized parameters used by the model for binary classes are introduced into the multiclass model containing all the classes. The algorithm is iterated for the multiclass model with the optimized parameters.

Since the model is trained with the training image dataset, it (the training image dataset) should fit the model accurately upon validation. Fig. 4 (a) shows that this model has 97% accuracy when validated with the training image dataset. Subsequently, if the training image dataset of only one of the classes fed to the algorithm is correct, the accuracy of the model is stunted to 1/7 (refer to Fig. 4(b)). Thus, the model predicts appropriately for the trained image dataset. The test image data set is unknown to the trained model. Thus, the model is validated using the test image data set to mimic a real-life scenario. Upon validation, the precision and recall values range from 88 to 99% and 81 to 97%, respectively, for different classes (see table 2). The overall accuracy of the multiclass model achieved is 92 % (see table 2). The overall accuracy of the model is calculated by averaging the values of precision of the individual classes through their equal shared weights. The loss curve in Fig. 5(b) signifies the reduction of signal noises as the training proceeds.

**Table 2:** Accuracy, precision, and recall values of a multiclass model consisting of 7 classes.

| Class | Accuracy | Precision | Recall |
| --- | --- | --- | --- |
| EA | 0.92 | 0.97 | 0.96 |
| SA |  | 0.94 | 0.99 |
| KP |  | 0.96 | 0.9 |
| AB |  | 0.91 | 0.88 |
| PA |  | 0.91 | 0.91 |
| ST |  | 0.95 | 0.99 |
| C |  | 0.81 | 0.98 |

**Figure 4.**
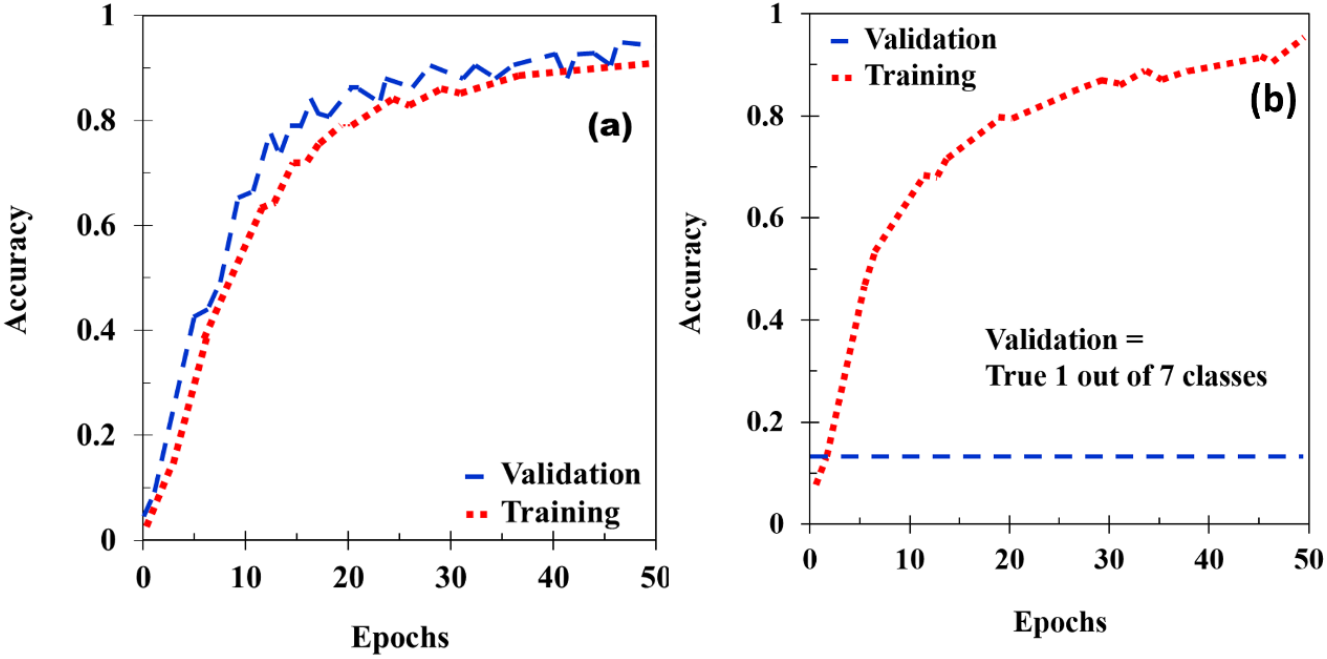
**Plots of validation curve for the multiclass model using training image dataset. (a) accuracy of the model validated using a training image set inclusive of all seven classes. (b) The accuracy of the model with only one dataset out of seven classes is correct. Thus the accuracy of the model is ∼1/7 (blue line in the graph).**

**Figure 5.**
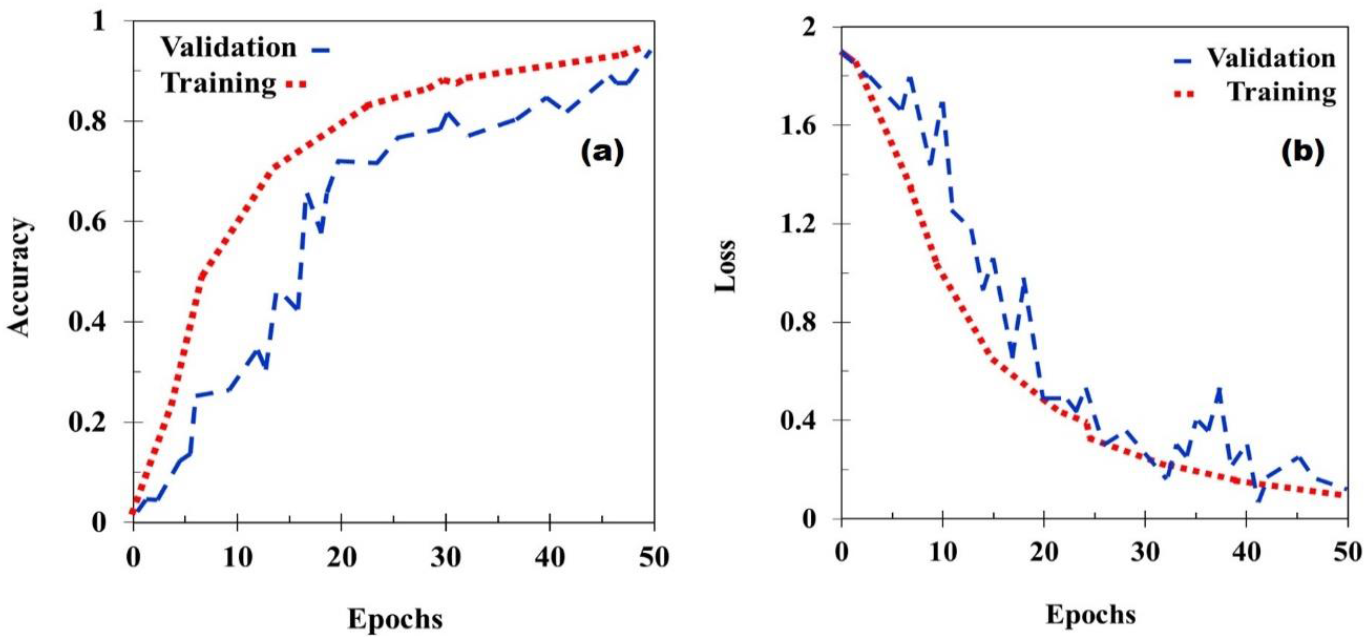
**Plots of (a) Accuracy curve and (b) loss curve of the multiclass model**

The precision of the multiclass model varies widely and can be classified based on its range. (1) The precision of class C is low <90%. (2) The precision of classes AB, PA, SA, and ST is between 90% to ≤95%. (3) EA and KP have precision >95%. The values of precision obtained in binary classes have a similar range for the given bacteria. For, e.g., binary classes of - EA and KP, PA and SA, and SA and ST have has 98%, 96%, and 98% precision, respectively. Therefore, it can be concluded that accuracy values are dependent on the individual bacteria and the extent of change they cause in the blood properties by their presence. Our investigation found that *Enterococcus faecium* (EF) (one of the ESKAPE pathogens) did not cause sufficient change in the blood properties, and the algorithm failed to distinguish the blood droplet residues that were infected with EF. Hence, the bacterial species EF has not been considered for investigation in the article.

In conclusion, we have demonstrated a technique to detect six bacterial species in the blood by recognizing the patterns found in the dried residue of blood droplets using CNN. The detection process is cost-efficient as it does not require any biomarkers/chemicals or high-end equipment. Since the detection procedure involves only capturing an optical image, this can be performed by a non-professional with minimal expertise in the healthcare sector. The trained algorithm developed into a mobile application with a user interface has the potential to revolutionize POC diagnostics. Moreover, the promising results with high accuracy obtained in this article are equivalent to the accuracy of standard bacteriological tests^57–59^. Besides, the technique has added benefits such as rapid turnaround time (less than 15 minutes), low cost, and is suitable for POC.

## EXPERIMENTAL PROCEDURE

All bacterial cultures, namely, *Enterobacter aerogenes, Staphylococcus aureus, Klebsiella pneumoniae, Acinetobacter baumannii, Pseudomonas aeruginosa, and Salmonella Typhi*, were grown in Luria Bertani medium. Briefly, a single colony of bacterial culture from a freshly streaked LB agar plate was inoculated in LB broth overnight at 37°C at 170 *rpm* shaker incubator. Next, the bacteria are normalized with OD_600nm_. The blood was collected in a lavender vacutainer (K3-EDTA, Quantum Biomedicals) from five healthy volunteers, age 24-30 years, with diverse blood groups (O +ve, O -ve, AB +ve, B +ve, B -ve), and it was stored at 4°C and used for experiments up to a week. These storage conditions do not influence the blood properties (for a week), and thus the drying process is unaffected due to storge^36^. Bloodstream infection diagnosis ranges from 1-10 CFU/mL^60,61^ to 10^3^-10^4^ CFU/mL^62^; blood collected from the donor was thus spiked with bacterial culture to a final concentration of 10^3^ CFU/mL to mimic tangible infection in real-life patients, and un-spiked blood is used as a control (C). Since equal volumes of phosphate-buffered saline (PBS-137 mM NaCl, 2.7 mM KCl, 8 mM Na_2_HPO_4_, and 2 mM KH_2_PO_4_) is added to normalize all the bacterial numbers to a constant of 1000 CFU/mL, an equal volume of PBS without any bacteria was added as a control. To observe bacterial-induced changes in the blood (as per the model of bloodstream infection), we incubated the blood at static conditions 37°C for 24 hours^64^ before drop-casting it. Subsequently, the control sample (C) is also placed in the same static conditions for 24 hours before drop-casting the same.

### Experimental setup for droplet evaporation and imaging

Optically flat plain glass slides procured from Blue Star© are cleaned using the following procedure: the glass slides are immersed in an ultrasound bath sonicator (from Rivotek^™^) containing isopropyl alcohol (commonly known as isopropanol) and are sonicated for 15 minutes. This is followed by a de-ionized (DI) water rinse and blow-dry. The average surface roughness (R_a_) is ∼40 nm (measured using the optical profilometer tool by The TalySurf CCI). As shown in Figures 1 (b) and (c), the incubated samples from the vials are drop cast gently onto the clean glass slides using a micropipette (Finnpipette®). The volume of the droplets ranges from 0.5 to 2 μl. Hundreds of droplets are placed on the glass slide in linear arrangements separated at least by the distance of 2 times the droplet diameter (*L*_*C*_), ensuring no vapor-mediated interactions between the individual droplets^65^ (refer to Fig.1 (c)). The glass slides are labeled as the sample deposited on a particular slide is known. The droplets are allowed to evaporate under controlled atmospheric conditions (26 ±5 °C and 45 ±10% relative humidity measured by TSP-01, Thorlabs) in an acrylic enclosure to minimize the external convection disturbances. All experiments are conducted inside a bio-safety hood with HEPA filters following the biosafety laboratory -II protocols. The bacterial samples and other plastic wares were discarded with 10% bleach, followed by double autoclaving all the waste.

The blood drops generally evaporate in 10-12 minutes; they are imaged using an optical microscope (Olympus microscope). The droplet is illuminated by an LED light source in-line with the objective lens of the microscope—a digital camera (Nikon D7200) attached to the microscope captures the reflected light and images of the droplet. The captured images were manually filtered for uniformity and shape (as the Bo<<1, the droplet assumes a spherical shape). The background of the images (other than the droplet region) is removed to reduce the redundant information. The captured images constitute the image data set to be processed through the CNN algorithm.

## Credit statement

Conceptualization: SB, DC; Methodology: OH, RC, DR; Investigation: OH, RC, VJ, Visualization: OH, RC, DR, VJ; Funding acquisition: SB, DC; Project administration: OH, VJ; Supervision: SB, DC; Writing original draft: OH; Editing and revision: OH, RC, VJ, DC, SB.

## Data Availability

All relevant data are within the paper and its Supporting Information files. All materials and additional data are available from the corresponding author upon request.

## Acknowledgments

The authors acknowledge Suraksha Sunil, Amey Agharkar, Ankur Chattopadhyay, Srinivas Rao S, Rovin Pinto, and Gannena K.S. Raghuram for assistance in drop-casting and imaging of blood droplets. The authors acknowledge all the volunteers who participated in blood donation - Amey, Anmol, Vivek, Omkar, Durbar. SB: DRDO Chair Professorship, DC: Infrastructure support from ICMR (Center for Advanced Study in Molecular Medicine), DST (FIST), UGC-CAS (special assistance), DAE-SRC fellowship, ASTRA-Chair fellowship, TATA Innovation grant, DBT-IOE partnership grant. RC duly acknowledges CSIR-SRF for financial assistance.

## Ethics declarations and approval

Human Ethical Review Committee of the Indian Institute of Science, Bangalore, approved the experimental protocols.

## Informed consent

Informed consent was obtained from all participants/volunteers.

---

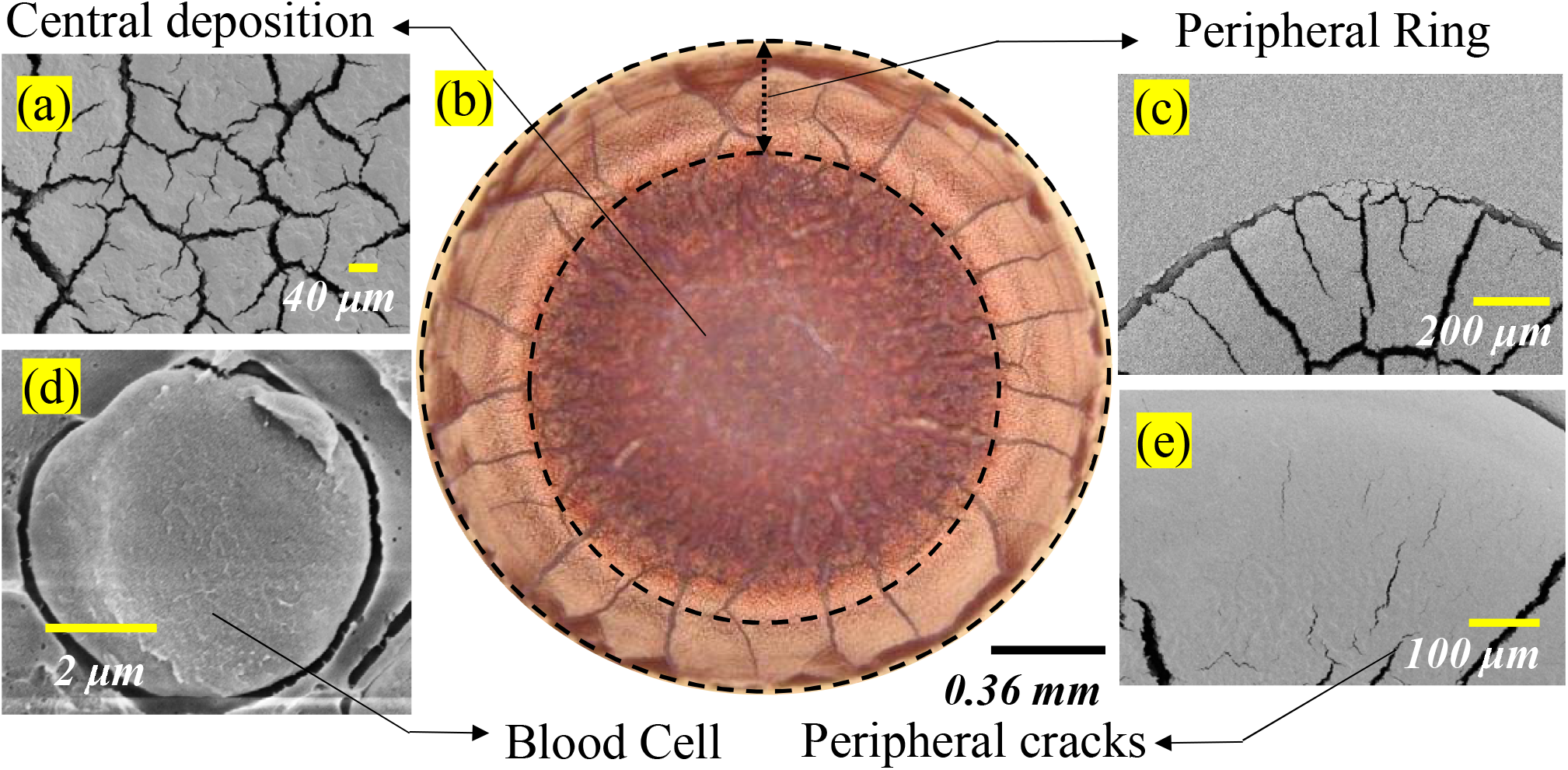

