## Supplementary Figure for "Convolution Neural Networks for Point-of-Care Diagnostics of Bacterial Infections in Blood"

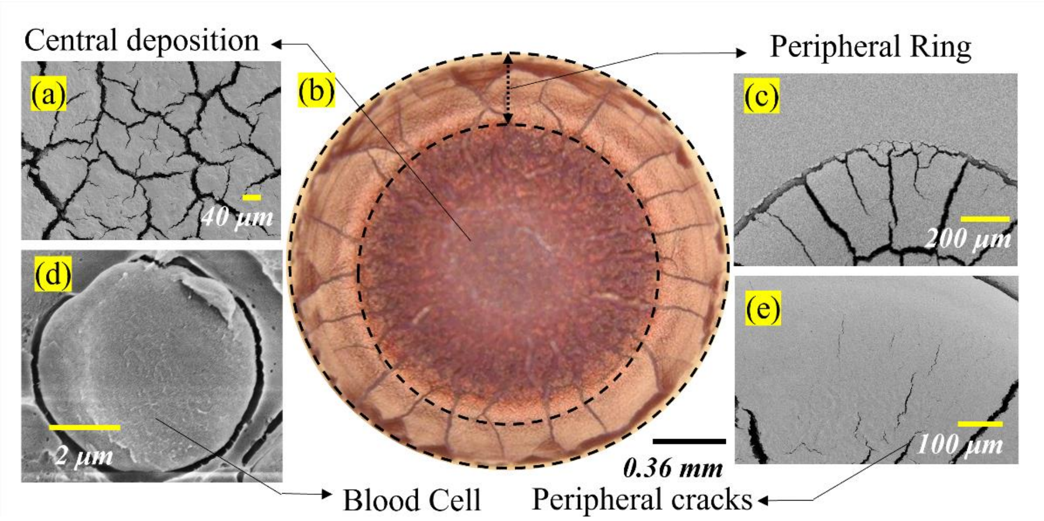


**S1: Dried image of a blood droplet from a healthy volunteer. (a) SEM image of the crack pattern at the center of the image. (b) Microscopic optical image of the entire blood droplet. SEM image of (c) crack patterns on the periphery of the droplet, (d) Red blood cell (RBC), (e) zoomed-in image of peripheral cracks on the dried residue.**
